# Detection of *Mycobacterium tuberculosis* transrenal DNA in urine samples among adult patients in Peru

**DOI:** 10.1101/2023.07.26.23293199

**Authors:** Annelies W Mesman, Roger I Calderon, Laura Hauns, Nira R Pollock, Milagros Mendoza, Rebecca C Holmberg, Molly F Franke

## Abstract

Diagnosis of tuberculosis (TB) relies on a sputum sample, which cannot be obtained from all symptomatic patients. *Mycobacterium tuberculosis (Mtb)* transrenal DNA (trDNA) has been detected in urine, an easily obtainable, noninvasive, alternative sample type. However, reported sensitivities have been variable and likely depend on collection/assay procedures and aspects of trDNA biology. We analyzed three serial urine samples from each of 75 adults with culture-confirmed pulmonary TB disease in Lima, Peru for detection of trDNA using short-fragment real-time PCR. Additionally, we examined host, urine, and sampling factors associated with detection. Overall sample sensitivity was 38% (95% Confidence Interval [CI] 30–45%). On a patient level (i.e., any of three samples positive), sensitivity was 73% (95% CI: 62-83%). Sensitivity was highest among samples from patients with smear-positive TB, 92% (95% CI: 62-100%). Specificity from a single sample from each of 10 healthy controls was 100% (95% CI: 69-100%). Adjusting our assay positivity threshold increased patient-level sensitivity to 88% (95% CI: 78-94%) overall without affecting the specificity. We did not find associations between *Mtb* trDNA detection and either patient characteristics or urine sample characteristics. Overall, our results support the potential of trDNA detection for TB diagnosis.

## Background

Tuberculosis (TB) remains a major global health burden, with an estimated 10.6 million cases and 1.6 million deaths in 2021.^1^ A major driver of this high mortality rate is delay in timely diagnosis. Therefore, there is an urgent need for diagnostic tools that can reliably diagnose TB in patients with culture-negative disease or who are unable to reliably expectorate sputum, such as children or people living with HIV.^2–7^

Urine is an appealing alternative sample to sputum because collection is non-invasive and produces no aerosols, thereby reducing transmission risk to health care workers. The currently available urine test, the lateral flow TB-LAM test, which detects presence of an *Mycobacterium tuberculosis (Mtb)* antigen, has limited specificity and sensitivity in and is only recommended for use among individuals living with HIV in inpatient settings.^8–11^ Diagnostics based on cell-free *Mtb* DNA released from infected apoptotic cells, known as transrenal (tr) DNA when it passes through the kidneys into urine, have demonstrated promise^12–15^ and do not rely on the presence of sputum. In spite of a strong theoretical basis for cell-free DNA-based diagnostics, test sensitivities in trDNA detection studies have varied enormously,^12–14,16–21^ likely reflecting a lack of understanding of trDNA biology and factors influencing presence of *Mtb* DNA in urine. Furthermore, some assays used for *Mtb* DNA detection, such as Xpert MTB/RIF (Cepheid, Sunnyville CA), are not designed to pick up the small DNA fragments that could be present in urine.^19–21^

The objective of this study was to report detection of *Mtb* trDNA in adults with culture-confirmed pulmonary TB disease using short-fragment real-time PCR and examine whether host, urine, or sampling factors could explain variability in *Mtb* trDNA detection.

## Methods

### Study population

As part of a larger diagnostic study, we recruited adults, at least 18 years of age, that were diagnosed with pulmonary TB disease through the National Tuberculosis Program in Lima, Peru and ultimately included only those who were culture-confirmed.^22,23^ For research purposes, all patients provided a sputum sample prior to TB treatment start (i.e. pre-treatment) for smear analysis, culture, and drug-sensitivity testing. Additionally, study staff collected data from the patient’s medical chart (history of prior TB treatment, diabetes, and chest radiography results) and conducted voluntary HIV testing.

### Urine sample collection

From each enrolled patient diagnosed with TB, we aimed to collect three urine samples. A first sample of urine was collected prior to treatment initiation and two additional samples were collected leveraging protocolized study visits occurring during the first week of treatment (days 4 and 7). Upon collection of urine (50 mL, clean catch), 5mL was separate for urinalysis and EDTA was immediately added (10 mM final) to the remaining sample to prevent nucleic acid degradation. Samples were transported via cold chain, aliquoted, and stored at -80°C. One aliquot from each sample was sent for urinalysis. All samples were collected between 6 and 11 AM in the morning. Urine samples from healthy individuals (one per person) were purchased from bioIVT (bioIVT.com) and similarly treated with 10mM EDTA.

### Laboratory procedures

For DNA extraction we used an anion exchange method, optimized for purification of low molecular weight nucleic acids, which was based on a protocol to detect fetal DNA in maternal urine. ^24^ In brief, we captured cell-free DNA in 20 mL of urine on Q-sepharose resin. The bound DNA was rinsed in a low-salt solution (0.3 M LiCl, 10 mM NaOAc) to remove PCR inhibitors before elution in a high salt (2 M LiCl, 10 mM NaOAc) buffer. Next, the sample was desalted and concentrated using a QIAquick spin column (Qiagen) in two washing steps (2 M LiCl in 70% EtOH, followed by 75 mM KoAC pH 5.0 in 80% EtOH) and eluted in a low salt buffer (QIAquick elution buffer EB; 50 μL).

We quantified *Mtb* trDNA via real time semi-nested PCR designed to detect a 39 base pair target from the IS6110 repeat following a protocol described by Shekhtman et al. ^24^ The reaction was set up with Jumpstart Taq (Sigma) using 200 nM of first stage forward primer, 700 nM of the second stage forward and reverse primer and 200 nM of the Taqman probe. We used 5 μL of urinary total DNA for quantification in a Roche Lightcycler 480. Samples were tested in duplicate. In every run we used a 1:1000 dilution in sterile urine of *Mtb* H37Ra cells (0.5 McFarland standard) for the positive control. The negative control was sterile urine.

### Data analysis

We calculated sensitivity and specificity and 95% confidence intervals (CI) both per sample (adjusting CIs for clustering by patient), and per patient. We estimated sensitivity overall and stratified by smear status and time of collection relative to treatment start. Specificity was estimated in ten commercially obtained urine samples from healthy individuals.

We considered samples with a Crossing Point (Cp) value < 35 and fluorescence increase > 10 units within 35 cycles as positive. This 10 units threshold was established using extracted TB-negative sputum spiked with *Mtb* genomic DNA. Additionally we conducted sensitivity analyses in which we lowered the positivity threshold for fluorescence to an increase of > 5 units. We also examined within-person change in Cp and fluorescence between baseline and day 4 and baseline and day 7. A decrease in Cp value and/or an increase in fluorescence between baseline and subsequent samples would indicate a greater quantity of *Mtb* trDNA shed in later samples.

To examine associations between *Mtb* detection in urine samples and factors related to sample collection (i.e., time of collection relative to TB treatment initiation), disease burden (i.e., smear positivity), comorbidity (i.e., diabetes mellitus) and urinalysis results (i.e. first urine of day, specific density, presence of bacteria, protein or yeast, pH, presence of leukocytes or red blood cells), we conducted binomial generalized estimating equation (GEE) regression analyses. We adjusted analyses for clustering by patient using an unstructured correlation structure. Statistical analyses were conducted in SAS Studio (Version onDemand for Academics SAS Institute Inc., Cary, NC, USA).

## Results

### Study population

We included 75 patients who had ≥ two urine samples collected, including a pre-treatment sample. Most patients (n=71, 95%) had three samples collected. Patient characteristics are presented in Table 1. The median age among patients was 27 years (range: 18-76) and 39% (n=29) were female (Table 1). Nineteen patients (25%) had a smear-negative sputum sample; 56 patients (75%) had a smear-positive sputum sample (Table 1). None of the patient were diagnosed with extrapulmonary TB disease. Two patients (3%) were living with HIV, and two patients (3%) self-reported diabetes.

**Table 1.** Characteristics of adult patients with sputum culture-positive pulmonary tuberculosis (N=75)

| Characteristic | n (%) |
| --- | --- |
| Age, median (Q1-Q3) | 27 (21-34) |
| Female sex | 29 (39) |
| Smear grade |  |
| Negative | 19 (25) |
| Positive 1+ | 22 (29) |
| Positive 2+ | 22 (29) |
| Positive 3+ | 12 (16) |
| Multidrug-resistant tuberculosis* | 10 (14) |
| Diabetes | 2 (3) |
| HIV* | 2 (3) |

\*Drug susceptibility test results and HIV status were missing for two patients

### Sample-level detection of trDNA

Overall assay sensitivity among all patient samples was 38% (95% CI: 30-45%) and was similar across collection times (i.e., baseline and 4 and 7 days post-treatment; Table 2). At each timepoint, sensitivity was lowest among samples collected from patients with smear-negative TB (26% [95% CI: 5-48%], 24% [95% CI: 1-46%], 37% [95% CI: 13-61%] respectively). Sensitivity was highest among samples collected pre-treatment from patients with a high (3+) smear grade (50% [95% CI: 17-83%].

**Table 2.** Sample-level sensitivity of *Mtb* trDNA detection at each sample collection point; CI: confidence interval.

|  | All samples* |  | Pre-treatment |  | Day 4 post treatment |  | Day 7 post treatment |  |
| --- | --- | --- | --- | --- | --- | --- | --- | --- |
| Smear grade | n/N | Sensitivity (95% CI) | n/N | Sensitivity (95% CI) | n/N | Sensitivity (95% CI) | n/N | Sensitivity (95% CI) |
| Negative | 16/55 | 29 (13-45) | 5/19 | 26 (5-48) | 4/17 | 24 (1-46) | 7/19 | 37 (13-61) |
| Positive 1+ | 26/66 | 39 (25-54) | 7/22 | 32 (11-53) | 10/22 | 45 (23-68) | 9/22 | 41 (19-63) |
| Positive 2+ | 25/66 | 38 (25-51) | 10/22 | 45 (23-68) | 10/22 | 45 (23-68) | 5/22 | 23 (4-42) |
| Positive 3+ | 16/34 | 47 (31-63) | 6/12 | 50 (17-83) | 5/11 | 45 (10-81) | 5/11 | 45 (10-81) |
| All smear grades | 83/221 | 38 (30-45) | 28/75 | 37 (26-49) | 29/72 | 40 (29-52) | 26/74 | 35 (24-46) |

\*Confidence intervals adjusted for clustering per patient

Changing the fluorescence sample positivity threshold to an increase of > 5 units (with Cp < 35) increased overall assay sensitivity among all samples to 58% (95% CI: 51-66%; Suppl Table 1). At each timepoint, sensitivity was higher with this positivity threshold, ranging from 41% to 53% in patients with smear-negative TB and 50% to 68% (Suppl Table1a) in those with smear-positive TB.

### Patient-level detection of trDNA

On a patient level, sensitivity (determined by positivity of at least one of a patient’s samples) was 73% (95% CI: 62-83%) among the 75 patients (Table 3). Sensitivity was lowest among patients with smear-negative TB at 58% (95% CI: 36-80%) and highest among patients with a high (3+) smear grade at 92% (95% CI: 62-100%). Eleven percent of patients (95% CI: 5-20%) had all three samples positive (Table 3). When we changed the sensitivity threshold to > 5 units, overall sensitivity was 88% (95% CI: 78-94%); 79% (95% CI 54-94%) among patients with smear-negative TB and 92% (95% CI 62-100%) among patients with a high (3+) smear grade. We did not observe notable within-person changes in either Cp values or fluorescence difference, relative to baseline (Suppl Table 2).

**Table 3.**
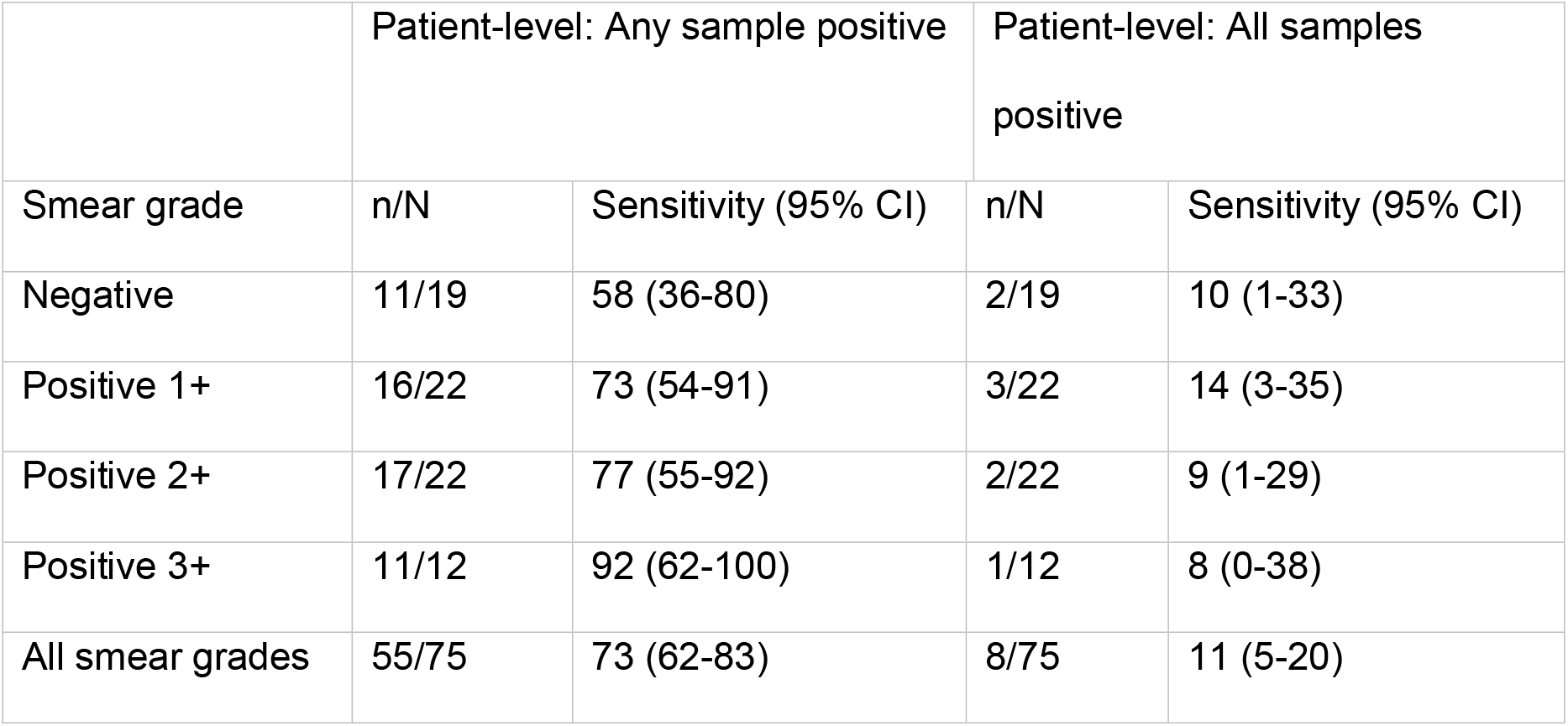
Patient-level sensitivity of *Mtb* trDNA detection, determined by any or all of a patient’s samples being positive.

|  | Patient-level: Any sample positive |  | Patient-level: All samples positive |  |
| --- | --- | --- | --- | --- |
| Smear grade | n/N | Sensitivity (95% CI) | n/N | Sensitivity (95% CI) |
| Negative | 11/19 | 58 (36-80) | 2/19 | 10 (1-33) |
| Positive 1+ | 16/22 | 73 (54-91) | 3/22 | 14 (3-35) |
| Positive 2+ | 17/22 | 77 (55-92) | 2/22 | 9 (1-29) |
| Positive 3+ | 11/12 | 92 (62-100) | 1/12 | 8 (0-38) |
| All smear grades | 55/75 | 73 (62-83) | 8/75 | 11 (5-20) |

### Specificity among controls

We observed a specificity of 100% (95% CI: 69-100%) based on a single analysis of ten commercially-obtained urine control samples. None of the samples tested positive. Specificity was not affected by changing the sample positivity threshold to an increase of > 5 units of fluorescence.

### Factors associated with Mtb trDNA detection in TB patients

We did not find associations between *Mtb* trDNA detection in urine samples and either patient-level characteristics or urine-sample characteristics (Table 4).

**Table 4.** Sample-level analysis of factors associated with *Mtb* trDNA detection in urine samples from patients with pulmonary TB disease.

|  | Positive signal | Negative signal<sup>#</sup> | p-value<sup>*</sup> |
| --- | --- | --- | --- |
|  | N=83<br>n (%) | N=138<br>n (%) |  |
| Female sex | 35 (42) | 52 (38) | 0.54 |
| Age (years) |  |  | 0.51 |
| <22 | 29 (40) | 33 (24) |  |
| 22-30 | 32 (39) | 46 (33) |  |
| 31-40 | 12 (14) | 33 (24) |  |
| >40 | 10 (12) | 26 (19) |  |
| Smear grade |  |  | 0.34 |
| Negative | 16 (19) | 39 (28) |  |
| Positive 1+ | 26 (31) | 40 (29) |  |
| Positive 2+ | 25 (30) | 41 (30) |  |
| Positive 3+ | 16 (19) | 18 (13) |  |
| Multidrug-resistant tuberculosis<sup>§</sup> | 12 (15) | 18 (13) | 0.68 |
| Diabetes | 1 (1.2) | 5 (3.6) | 0.25 |
| Urinalysis results |  |  |  |
| First urine of the day<sup>†</sup> | 15 (18) | 31 (23) | 0.46 |
| Specific density |  |  |  |
| <1.01 | 16 (33) | 32 (23) | - |
| 1.01-1.025 | 67 (40) | 101 (60) |  |
| >1.025 | 0 (0) | 4 (3) |  |
| Bacteria | 4 (4.8) | 14 (10) | 0.07 |
| Protein | 1 (1.2) | 1 (0.7) | 0.49 |
| Yeast | 3 (3.6) | 3 (2.2) | 0.46 |
| pH |  |  | 0.56 |
| Acid | 78 (94) | 130 (95) |  |
| Neutral | 4 (4.8) | 4 (2.9) |  |
| Alkaline | 1 (1.2) | 3 (2.2) |  |
| Leukocytes | 20 (24) | 33 (24) | 0.82 |
| Red blood cells | 18 (22) | 31 (23) | 0.94 |

## Discussion

In a cohort of adult patients with culture-confirmed pulmonary TB disease, 73% had trDNA detected in at least one of three urine samples, with sensitivity increasing with each additional sample collected. While the patient-level sensitivity observed in our study is higher than a study in South Africa that found 42% sensitivity among patients with culture-confirmed TB disease ^13^, it falls just below that reported in two other studies that detected trDNA in 79% and 88% of culture-confirmed TB patients.^12,14^ Notably, we found a higher sensitivity with higher smear grades, a trend that has been repeatedly observed for non-sputum based diagnostic approaches.^25–28^ Varying sensitivity by smear grade has implications for drawing comparisons across studies, in that, if sputum bacillary load is an important determinant of test sensitivity and this varies across population, average diagnostic performance would be expected to vary by population as well. Importantly, low sensitivity among individuals with the lowest bacillary burden highlights the need to optimize trDNA purification and/or detection assay protocols to maximize sensitivity among patients with a low bacillary load. A recent study used a CRISPR-based approach detect cell free *Mtb* DNA in serum samples, including from children and unconfirmed clinically-diagnosed TB cases.^29^ This promising technique may also work to amplify the signal in urine samples.

We did not identify patient-level characteristics, urine-related factors (i.e. pH, presence of blood cells or protein) or sampling factors (i.e., first void of day, collection after treatment initiation) that impacted sensitivity. This suggests that, at least in this cohort, these factors are not major drivers of test positivity. While sensitivity increased with increasing sputum smear grade, very few patients had a positive result on all three of their urine sampling, highlighting within-person variability that was not explained by the factors we examined. Several studies have found higher sensitivities for trDNA assays among patients living with HIV and with extrapulmonary TB;^14,16^ however, it is possible that these higher sensitivities were attributable to disseminated TB disease in these patients. Differences in procedures across studies, whether in sample collection or trDNA capture or detection could make it challenging to directly compare across studies.^12–14^

Limitations of this initial study include that we did not include patients for whom this method could be most relevant (i.e. patients with culture-negative TB, living with HIV and children) and only included urine samples from healthy donors as control group.

## Conclusions

In summary, these results reinforce the feasibility of *trDNA* as a promising diagnostic approach and highlight bacillary burden as a major driver of sensitivity. The patient, urine, and sampling characteristics that we examined did not contribute importantly to variation in detection with this assay.

## Data Availability

The datasets used and/or analysed during the current study are available from the corresponding author on reasonable request.

## List of Abbreviations

(TB): tuberculosis
(trDNA): transrenal DNA
(Mtb): *Mycobacterium tuberculosis*

## Ethics approval and consent to participate

Study participants provided written informed consent. All study procedures were approved by the Ethics Committee of Peru’s National Institute of Health (OEE-039-14) and the Office of Human Research Administration at Harvard TH Chan School of Public Health (IRB 13-2754).

### Consent for publication

*Not Applicable*

### Funding

Research reported in this publication was entirely supported by *the National Institutes of* Allergy and Infectious Disease of the National Institutes of Health under award numbers U19AI109755 and R03AI153554. The content is solely the responsibility of the authors and does not necessarily represent the official views of the National Institutes of Health.

### Authors’ contributions

AWM: Data analysis; First draft of the paper

RIC: Scientific leadership in laboratory procedures; Data interpretation

LH: Led and implemented laboratory procedures

NRP: Scientific leadership in laboratory procedures; Data interpretation

MM: Acquisition of clinical samples and data

RCH: Scientific leadership in laboratory procedures; Data interpretation

MFF: Conceived of the study; substantively revised the manuscript

All authors read and approved the final manuscript.

## Acknowledgements

We are indebted to the DETECT-TB study staff at Socios En Salud, to the individuals who chose to participate in this study, and to the Peru National Tuberculosis Program staff who supported this work.

---

**Suppl Table 1a:**
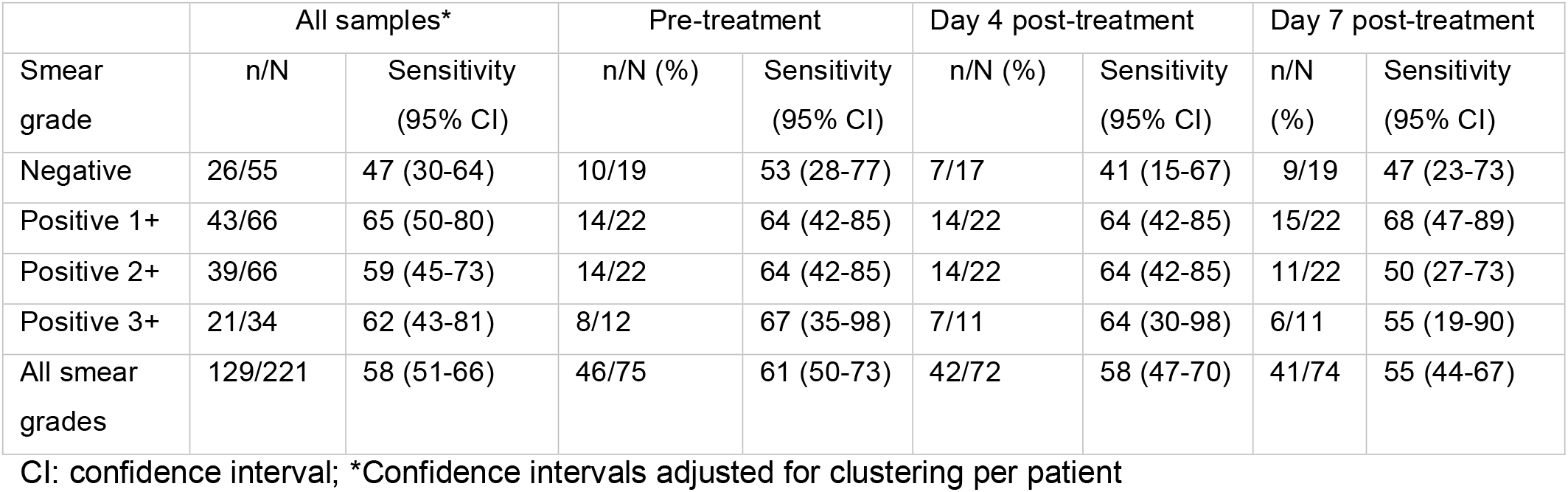
Sample-level sensitivity of *Mtb* trDNA detection at each timepoint applying a fluorescence threshold > 5.

**Suppl Table 1b.** Patient-level sensitivity of *Mtb* trDNA detection, determined by any or all of patient’s samples having a fluorescence threshold > 5.

|  | Patient-level: Any of (three) samples |  | Patient-level: All of (three) samples |  |
| --- | --- | --- | --- | --- |
| Smear grade | n/N | Sensitivity (95% CI) | n/N | Sensitivity (95% CI) |
| Negative | 15/19 | 79 (54-94) | 3/19 | 16 (3-40) |
| Positive 1+ | 20/22 | 91 (79-100) | 8/22 | 36 (17-59) |
| Positive 2+ | 20/22 | 91 (79-100) | 6/22 | 27 (11-50) |
| Positive 3+ | 11/12 | 92 (62-100) | 3/12 | 25 (5-57) |
| All smear grades | 66/75 | 88 (78-94) | 20/75 | 27 (17-37) |

CI: confidence interval.

**Suppl Table 2.** Median Cp and fluorescence values at each timepoint, by smear grade.

|  | Cp |  |  | Fluorescence |  |  |
| --- | --- | --- | --- | --- | --- | --- |
| Smear grade | pre-tx | day 4 post-treatment | day 7 post-treatment | pre-tx | day 4 post-treatment | day 7 post-treatment |
| Negative | 34.0 | 35.0 | 34.0 | 5.2 | 2.9 | 2.4 |
| Positive 1+ | 33.5 | 31.5 | 32.2 | 7.9 | 9.5 | 8.3 |
| Positive 2+ | 32.3 | 33.2 | 33.8 | 9.1 | 8.9 | 5.1 |
| Positive 3+ | 32.2 | 34.1 | 30.7 | 10.2 | 7.3 | 8.6 |
| All smear grades | 33.2 | 33.6 | 33.1 | 6.3 | 8.1 | 7.4 |

tx: treatment

